# Does the sensitivity- and precision-maximizing RCT filter find all ‘included’ records retrieved by the sensitivity-maximizing filter on Ovid MEDLINE? An investigation using 14 Cochrane reviews

**DOI:** 10.64898/2026.03.20.26348876

**Authors:** Helen Fulbright, David Marshall, Connor Evans, Mark Corbett

## Abstract

**Objectives:** To inform users about the impact of two updated study filters for limiting database search results to randomized controlled trials on Ovid MEDLINE: a sensitivity-maximizing version (SM) and a sensitivity-and-precision-maximizing version (SaPM). To provide an updated understanding of how they compare to each other.

**Methods:** Using the final ‘included’ records of 14 Cochrane reviews that had used the SM filter, we determined how many available records on Ovid MEDLINE would have been retrieved with each filter; investigated why records were missed; the unique yield; precision; and number-needed-to-read (NNR) for each filter. We also performed forwards and backwards citation searching on missed records (to determine if this could mitigate the risk of missing ‘includes’) and calculated the percentage change in the overall number-needed-to-screen (ONNS) when applying each filter to reproduction strategies.

**Results:** On average, the SaPM filter reduced ONNS by 83% and retrieved 95.9% of ‘includes’ compared with 98.2% retrieved by the SM filter. The SaPM filter offered a further 28.2% mean reduction in ONNS over the SM filter. The SM filter had a unique yield of 12 and a precision of 1.5%, versus a unique yield of three and precision of 4.4% for the SaPM filter. NNR was 68 for the SaPM filter versus 189 for the SM filter.

**Conclusion:** The SaPM filter reduced the screening burden with minimal risk of missing eligible records (which could be mitigated by citation searching). Decisions about which filter to use should consider both the needs and resources of the review.

## INTRODUCTION

With the volume of literature on MEDLINE growing daily, it is important to maintain a sensitive and systematic approach to literature searching while ensuring a manageable volume of records for review teams. A study filter is a pre-made search strategy designed to retrieve results for a specified study design [1] which can be used reduce the number of irrelevant records and the overall screening burden. However, it can be difficult to assess which filter to use for individual evidence synthesis projects, especially when there are many to choose from.

In 1994, Lefebvre developed the Cochrane highly-sensitive search strategy to help identify randomized trials in MEDLINE records [2]. There are now two validated highly-sensitive search strategies available for MEDLINE via the Ovid platform: a sensitivity-maximizing version (SM) [3] and a sensitivity-and-precision-maximizing version (SaPM). Both filters are periodically updated and can be found in the Technical Supplement to Chapter 4 in the *Cochrane Handbook for Systematic Reviews of Interventions* [4].

Updates to the filters in 2008 were tested in a 2020 study by Glanville et al, comparing 38 different randomized controlled trial (RCT) filters on Ovid MEDLINE [5]. This study reported 0.96 sensitivity and 0.14 precision for the SM filter versus 0.93 sensitivity and 0.46 precision for the SaPM filter [5]. Cochrane recommends using the SaPM filter, only if the number of records retrieved by the SM filter is unmanageable [3]. This means the SM filter is routinely used in evidence syntheses, suggesting the value of understanding the SaPM filter’s performance in relation to it. The SaPM filter could reduce the screening burden and save time for larger review topics or rapid reviews, where the timeliness and efficiency of review processes may be particularly important. The updated SM and SaPM filters have not yet been compared. The SM and SaPM filters (see Table 1) have an overlapping design but differ in their use of certain subject headings, subheadings, search terms and field codes. The update to both filters in 2023 exploded the subject heading Randomized Controlled Trial/ to retrieve records with the subject headings Equivalence Trial/ and Pragmatic Clinical Trial/, which were created in 2018 and 2014, respectively [6].

**Table 1:** Comparison of the 2023 updated RCT filters for MEDLINE.

| Line Number | MEDLINE Sensitivity-Maximizing Version (2023 Revision) | MEDLINE Sensitivity- and Precision-Maximizing Version (2023 Revision) |
| --- | --- | --- |
| 1 | exp randomized controlled trial/ | exp randomized controlled trial/ |
| 2 | controlled clinical trial.pt. | controlled clinical trial.pt. |
| 3 | randomized.ab. | randomized.ab. |
| 4 | placebo.ab. | placebo.ab. |
| 5 | **drug therapy.fs.** | **clinical trials as topic.sh.** |
| 6 | randomly.ab. | randomly.ab. |
| 7 | **trial.ab.** | **trial.ti.** |
| 8 | **groups.ab.** | or/1-7 |
| 9 | or/1-8 |  |

/ or **sh** = Medical Subject Heading; **ti** = title field; **ab** = abstract field; **dt** = drug therapy; **fs** = floating subheading; **pt** = publication type. **NB:** Differences between the two filters are shown in bold. Only the lines relevant to RCTs are shown, the limit to remove animal studies within the filters is not shown here.

Understanding how different study filters compare assists decision-making for information specialists. Since Cochrane recommend beginning with the SM version of the filter [3], it would be valuable to test the SaPM filter in the context of the SM filter’s performance using the results of published systematic reviews. It would be useful to explore how many records that meet the eligibility criteria after full-text screening (i.e., the final ‘includes’) retrieved by the SM filter would also be retrieved with the SaPM filter. Calculating differences in the precision, number-needed-to-read (NNR), and the overall number-needed-to-screen (ONNS) would allow comparison of the reduced screening burden offered by each filter. Precision refers to the proportion of relevant records retrieved out of the total number of records retrieved [3] and is typically low for systematic reviews with high sensitivity [7]. Within the context of systematic reviews, increases in precision are therefore most desirable where sensitivity remains high. This metric is a useful for comparing the ‘efficiency’ of filters in terms of the screening burden: as precision increases, the screening burden decreases. NNR is the number of records that need to be read on title and abstract to find one ‘include’ [8], with a lower NNR indicative of higher precision. But there is no ‘optimum’ threshold for NNR set for systematic reviews [9]. ONNS is the overall number of records that need to be screened on title and abstract. In addition to comparing these differences between the filters, it would be beneficial to examine if a particular filter’s disadvantages could be mitigated with citation searching. This is a supplementary search method which refers to the identification of studies that: cite a specified study of interest (forwards citation searching) or have been cited by the specified study (backward citation searching) [3].

The *Cochrane Handbook* advises that search filters should be assessed for their current accuracy and effectiveness due to regular updates to database interfaces and indexing [3]. However, updates to the filters in 2023 are not the only factor that might affect their performance in comparison to each other. One such change on MEDLINE since April 2022, is the machine-indexing of medical subject headings (MeSH) and publication types [10]. This has the potential to affect the ongoing performance of the filters if machine-indexing either: does not apply the RCT subject headings or publication types used within the filters when it should or applies these incorrectly. However, machine-indexing cannot affect the retrieval of any records using the free-text terms of each individual RCT filter, as these will continue to be retrieved based on their appearance in the search field specified. This is a further reason to provide an updated comparison of the filters.

## OBJECTIVES

This paper aims to understand how the updated SM and SaPM study filters compare to each other in limiting database search results to RCTs on Ovid MEDLINE. Using the results of 14 Cochrane reviews published or updated since April 2022, this study investigates: how many of the final ‘includes’ (i.e., records meeting the eligibility criteria after full-text screening) that are available on Ovid MEDLINE are retrieved by each filter; each filter’s unique yield; and why ‘includes’ were missed by each filter. We explore if any records missed by the filters can be found through forwards or backwards citation searching (FCS or BCS). In addition, the study recreates the search strategies of nine of the 14 Cochrane reviews to calculate the precision; NNR and the percentage change in the ONNS when applying each version of the filter.

## METHODS

A summary of the methodology is as follows, we:

1. found eligible datasets, composed of the ‘includes’ of 14 Cochrane reviews that limited records to RCTs using the SM filter;
2. determined how many of the ‘includes’ on Ovid MEDLINE would have been retrieved with each filter; investigated why records were missed (where applicable); and determined whether any records were retrieved with one filter only (unique yield);
3. performed forwards and backwards citation searching on records missed by the filters, to determine if the risks of missing ‘includes’ could be mitigated;
4. reproduced the search strategies for nine of the 14 Cochrane reviews and calculated the precision, NNR and the percentage change in the ONNS when applying each filter.

### 1. Finding eligible datasets

The study was conducted due to the Centre for Reviews and Dissemination’s (CRD) interest in the ‘risks’ and ‘benefits’ of using the SaPM filter over the SM filter. So that the filters could be tested on a range of review topics comparable to those worked on at CRD, we reviewed our publication list for 2019-2024 and created 61 single-line search terms for various conditions or populations appearing in the titles of studies. Search terms were designed to retrieve a range of studies on mental health conditions, public and population health, acute care, and various other diseases and chronic health conditions. Using Cochrane reviews provided us with access to a greater number of recent reviews on these topics than were available to us.

Using the 61 single-line search terms, two searches were performed on the Cochrane Database of Systematic Reviews (CDSR) via the Wiley platform on 2^nd^ January 2025 and 16^th^ April 2025. The searches were limited by publication date to find recently published or updated reviews. In total, we selected 14 Cochrane reviews, even though there were additional eligible reviews. This number was considered appropriate for a small-scale project and ensured we had more than 100 ‘includes’ overall, which Sampson suggests may be an appropriate number to use to validate methodological filters [11]. All search strategies are available in the appendix.

Reviews were considered eligible if they had: searched Ovid MEDLINE (either directly or through searches of specialized registers of trials and studies compiled by Cochrane); limited to RCTs using the SM version of the filter (either alone or in addition to adaptations to increase sensitivity); run or updated the searches in or after April 2022; and not used any date limits. Reviews using minor adaptations to the SM filter to enhance sensitivity were considered appropriate because this reflects typical practices employed by information specialists, and all records that would be retrieved with the SM filter would continue to be retrieved. This methodology means the performance of the SaPM filter is being evaluated in the context of records retrieved by the SM filter, rather than being a true comparison between the filters. Due to differences in the design of the filters, which mean they can each retrieve unique records, the SaPM filter has further potential to retrieve unique records than we can observe or report in this study.

After identifying 14 eligible reviews, we downloaded the final ‘includes’ for each Cochrane review (i.e., records meeting the eligibility criteria after full-text screening) into EndNote libraries, separating results into datasets for each review. Table 2 shows details of the datasets used, with the number of ‘included’ records that were found on Ovid MEDLINE ranging from 10 to 54. Further details about the datasets are available in the appendix.

**Table 2:** Datasets used.

| **Dataset:** | **Cochrane Review:** | **Searches Last Run:** | **Records ‘Included’ in Cochrane Review:** | **‘Included’ Records Found on MEDLINE:** | **SM Filter (2008 Revision) Adaptations & Additional Lines:** |
| --- | --- | --- | --- | --- | --- |
| 1 | Campisi et al. (2024) | May 2023 | 23 | 19 | randomized.ab. changed to randomi#ed.ab.<br>Additional lines included |
| 2 | Ernst et al. (2024) | October 2023 | 54 | 43 | randomized.ab. was changed to randomi?ed.ab.<br>Cooper et al's Phase III (P3) filter was used |
| 3 | Esezobor et al. (2024) | March 2023 | 33 | 23 | None |
| 4 | Kolkailah et al. (2024) | March 2023 | 53 | 24 | None |
| 5 | Kongwattanakul et al. (2024) | December 2022 | 70 | 43 | None |
| 6 | Sachdeva et al. (2024) | November 2024 | 13 | 12 | None |
| 7 | Ferraro et al. (2025) | November 2024 | 53 | 22 | None |
| 8 | Reid et al. (2025) | July 2023 | 20 | 14 | None |
| 9 | Wagner et al. (2025) | October 2023 | 59 | 54 | randomized.ab. was changed to randomi?ed.ab.<br>Cooper et al's Phase III (P3) filter was used |
| 10 | Chan et al. (2025) | March 2023 | 55 | 54 | randomized.ab. was changed to randomi?ed.tw. |
| 11 | Al Said et al. (2025) | October 2023 | 23 | 10 | None |
| 12 | Spillane et al. (2025) | March 2023 | 17 | 15 | None |
| 13 | Kreuzberger et al. (2025) | November 2023 | 138 | 49 | randomized.ab. was changed to randomi?ed.ab.<br>Cooper et al's Phase III (P3) filter was used |
| 14 | Wang et al. (2025) | January 2024 | 14 | 12 | randomized.ab. was changed to randomi*ed.ab. |

**Dataset citations:** Dataset 1 [12], Dataset 2 [13], Dataset 3 [14], Dataset 4 [15], Dataset 5 [16], Dataset 6 [17], Dataset 7 [18], Dataset 8 [19], Dataset 9 [20], Dataset 10 [21], Dataset 11 [22], Dataset 12 [23], Dataset 13 [24], Dataset 14 [25].

### 2. Recall and unique yield of SM and SaPM filters

We allocated the final ‘includes’ of each of the 14 Cochrane reviews to individual EndNote libraries and removed any duplicates. For each of these 14 datasets, we then created search strings to find which of these ‘includes’ were available on Ovid MEDLINE, so that we could report on each filter’s recall (also known as sensitivity). For this part of the study, we did not reproduce the original search strategies in case some of the ‘includes’ available on MEDLINE had been missed by the Cochrane review’s original MEDLINE strategy and found on other databases or sources used within the review.

Retrieving the ‘includes’ available on MEDLINE was achieved by searching for all or part of the title of the record as an exact phrase and checking if the result found matched the metadata of the record in the dataset. In some cases, additional data, such as an author surname, was used to limit the search results to the correct record. Records that did not appear to be available on MEDLINE were double-checked, using shorter sections of the study title that were unlikely to vary due to differences in spelling, punctuation, or special characters.

Once all ‘includes’ available on MEDLINE had been retrieved in the search strategy, we applied the SM and SaPM filters, with the final line of each filter individually combined with the line retrieving the ‘includes’. The ‘includes’ missed by each filter were then exported into EndNote groups labelled ‘Missed by SM filter’ or ‘Missed by SaPM filter’ for investigation. We checked whether either filter retrieved any unique records and exported these into EndNote groups labelled ‘Unique to SM filter’ or ‘Unique to SaPM filter’, accordingly.

In testing the filters, we did not include any lines to remove animal studies, as this section of the filter tends to be employed at various stages of a search (i.e., not always immediately following an RCT filter) and information specialists may not always wish to exclude these studies. MEDLINE(R) ALL was used for each dataset, regardless of the segment used in the original Cochrane review. Full strategies are available in the appendix.

For each dataset we calculated the recall (i.e., the number of ‘included’ records retrieved by a filter divided by the total number of ‘included’ records on MEDLINE) and unique yield (the number of unique records retrieved by one filter). Results using percentages are shown and rounded to one decimal place.

### 3. Citation searching of ‘includes’ missed by SM or SaPM filters

We used Web of Science and CitationChaser to perform forwards and backwards citation searching using the records missed by each filter for each dataset. This allowed us to establish if the risk of missing any ‘includes’ due to filter choice could be mitigated with citation searching,

Citation searching on Web of Science was conducted by entering the article titles of missed records and searching the title field, whereas digital object identifiers (DOIs) were used to search CitationChaser. We downloaded the forwards and backwards citations and labelled these in EndNote. For each missed record, we compared the forwards and backwards citations against the full list of ‘includes’ for the Cochrane review to see if any of these appeared in both sets. Finding a record in both the full list of ‘includes’ and either the forwards or backwards citations meant that the missed record would be found by citation searching.

Typically, the final ‘includes’ of a review would be used to perform citation searching [26]. However, for the purposes of this investigation it was more efficient to use the missed records, since there were fewer of them. This means that anything found backwards with the missed ‘includes’ would have been found forwards with the final ‘includes’ (and vice versa).

### 4. Precision, NNR and percentage change in ONNS

To report on the precision, NNR and percentage change in ONNS, we reproduced the original MEDLINE strategies for nine of the 14 Cochrane reviews and then applied the SM and SaPM filters. We excluded datasets that used a study filter on under 1,000 records as this is not representative of our typical use of the SM or SaPM filters. We also excluded datasets that had only searched specialized registers of trials and studies compiled by Cochrane, as this methodology is not representative of how CRD typically conducts systematic reviews. For these reasons, datasets 3, 5, 6, 7 and 11 were excluded from this part of the study.

All nine Cochrane reviews had used the 2008 version of the SM filter in their original MEDLINE strategies or had adjusted it to enhance sensitivity. The search strategies were therefore reproduced without any of the filters used originally, so that we could show the percentage change in ONNS when applying each updated version of the filter from 2023.

We applied date limits to remove records created or published after each Cochrane review’s searches were last updated. This allowed us to ensure our calculations for the precision (number of ‘included’ records retrieved by the MEDLINE strategy and retrieved with a filter, as a proportion of the total number of records retrieved with the filter applied to the search strategy) and NNR (1 divided by the precision) used a volume of records approximate to the number that would have been retrieved at the time the searches were run originally. It is important to note that the precision calculated for this study is therefore for each filter finding the ‘included’ records for each review using the reproduced MEDLINE strategies, and should not be confused with the precision for each filter finding RCTs. The value for NNR was rounded to the nearest whole number.

To calculate the percentage change in ONNS, we used the number of records retrieved by the reproduced strategy (without any filters), minus the number of records retrieved with the filter applied to the strategy, divided by the number of records retrieved by the strategy (without any filters), and multiplied this by 100. We also compared the percentage reduction of each filter and the differences in reduction volume between the filters.

All search strategies are available in the appendix. Results using percentages are shown and rounded to one decimal place.

## RESULTS

Table 3 shows the recall of both filters and the unique yield of each filter by each individual dataset. Totals are given for all datasets combined.

**Table 3:** Recall and unique yield.

| **Dataset:** | **Recall**<br>**(‘Includes’ Retrieved on MEDLINE)** |  | **Unique Yield**<br>**(Unique ‘Includes’ Retrieved)** |  |
| --- | --- | --- | --- | --- |
|  | **SM Filter:** | **SaPM Filter:** | **SM Filter:** | **SaPM Filter:** |
| 1 | 19 of 19 (100%) | 18 of 19 (94.7%) | 1 | 0 |
| 2 | 43 of 43 (100%) | 43 of 43 (100%) | 0 | 0 |
| 3 | 23 of 23 (100%) | 23 of 23 (100%) | 0 | 0 |
| 4 | 24 of 24 (100%) | 24 of 24 (100%) | 0 | 0 |
| 5 | 42 of 43 (97.7%) | 41 of 43 (95.3%) | 1 | 0 |
| 6 | 12 of 12 (100%) | 12 of 12 (100%) | 0 | 0 |
| 7 | 22 of 22 (100%) | 22 of 22 (100%) | 0 | 0 |
| 8 | 14 of 14 (100%) | 14 of 14 (100%) | 0 | 0 |
| 9 | 53 of 54 (98.1%) | 53 of 54 (98.1%) | 1 | 1 |
| 10 | 54 of 54 (100%) | 54 of 54 (100%) | 0 | 0 |
| 11 | 9 of 10 (90%) | 10 of 10 (100%) | 0 | 1 |
| 12 | 14 of 15 (93.3%) | 15 of 15 (100%) | 0 | 1 |
| 13 | 46 of 49 (93.9%) | 37 of 49 (75.5%) | 9 | 0 |
| 14 | 12 of 12 (100%) | 12 of 12 (100%) | 0 | 0 |
| **Total:** | **387 of 394 (98.2%)** | **378 of 394 (95.9%)** | **12** | **3** |

**SM** = Sensitivity-Maximizing Filter; **SaPM** = Sensitivity- and Precision-Maximizing Filter; **NB:** Grey cells are used to highlight a unique yield above 0.

For eight out of 14 datasets, the number of ‘includes’ retrieved was 100%, with no unique records found by either filter. However, the number of ‘includes’ retrieved was identical for nine out of 14 datasets overall, as recall was 98.1% for both filters in dataset 9 (with each filter also finding a unique record). The SM filter retrieved 12 unique records in total for all datasets versus three unique records retrieved by the SaPM filter. Overall, out of 394 records, the SM filter retrieved 387 (98.2%) MEDLINE ‘includes’ versus 378 (95.9%) by the SaPM filter.

Table 4 shows characteristics of the unique ‘includes’ that were missed for each filter and dataset. The SM filter missed seven records in total: three misses were unique to the filter with the remaining four missed by both filters. Records were missed by the SM filter and not the SaPM filter due to the inclusion of ‘trial’ in the title field (n=3) and ‘Clinical Trials as Topic’ as a subject heading (n=1). Two of the three unique missed records had no abstract.

**Table 4:** Characteristics of missing ‘includes’.

| **Filter:** | **Dataset:** | **Unique ‘Includes’ Missed:** | **Reason Found by Other Filter:** | **Alternative Filter Changes that would Retrieve Record:** | **Other Factors for Missed Record:** |
| --- | --- | --- | --- | --- | --- |
| **SM** | 9 | Broderick et al. (2014) | trial.ti.<br>Clinical Trials as Topic/ | group.ab. |  |
|  | 11 | Giustino et al. (2020) | trial.ti. | Randomized<br>Controlled Trials as<br>Topic/ | no abstract |
|  | 12 | Leyburn et al. (1959) | trial.ti. |  | no abstract |
| **SaPM** | 1 | McNamara et al. (2014) | trial.ab. |  | no subject headings |
|  | 5 | Raman et al. (1978) | groups.ab. |  | no subject headings |
|  | 9 | Toohey et al. (2018) | groups.ab. |  | no subject headings |
|  | 13 | Fiaccadori et al. (2023) | trial.ab. | trials.ti. |  |
|  |  | Barrington et al. (2016) | trial.ab.<br>dt.fs. |  |  |
|  |  | Eichenauer et al. (2021) | dt.fs. | randomized.ti. | no abstract |
|  |  | Kobe et al. (2018) | trial.ab.<br>dt.fs. | clinical trial, phase III.pt. |  |
|  |  | Marini et al. (2021) | trial.ab. |  | no subject headings |
|  |  | Mettler et al. (2018) | trial.ab. |  | no subject headings |
|  |  | Illidge et al. (2020) | dt.fs. |  | no abstract |
|  |  | Viviani et al. (2020) | trial.ab.<br>dt.fs. |  |  |
|  |  | Bari et al. (2020) | trial.ab. | clinical trial.pt.† |  |
| **Both Filters** | **Dataset:** | **Shared ‘Includes’ Missed:** | **Filter Changes that would Retrieve Record:** |  | **Other Factors for Missed Record:** |
|  | 5 | Lopez-Jaramillo et al. (1987) | (de and tu).fs. |  | no abstract |
|  | 13 | Oertel et al. (2023) | trials.ti,ab. |  | no subject headings |
|  |  | Zinzani et al. (2016) | clinical trial, phase II.pt.<br>phase II.ab. |  |  |
|  |  | Rigacci et al. (2020) | clinical trial.pt.<br>group.ab. |  |  |

**SM** = Sensitivity-Maximizing Filter; **SaPM** = Sensitivity- and Precision-Maximizing Filter; **/** = Medical Subject Heading; **ti** = title field; **ab** = abstract field; **dt** = drug therapy; **tu** = therapeutic use; **fs** = floating subheading; **pt** = publication type
†This may be worth using as a publication type for reviews unlikely to have many trials for the topic (e.g., uncommon diseases). However, it may be too sensitive for topics where there have been lots of trials (e.g., common cancers).

The SaPM filter missed 16 records in total: 12 misses were unique to the filter with the remaining four missed by both filters. A total of 12 records were missed by the SaPM filter and not the SM filter due to the inclusion of: ‘trial’ in the abstract field (n=8); drug therapy as a floating subheading (n=5); and groups in the abstract field (n=2). Two of these 12 unique records had no abstract and five had no subject headings.

‘Includes’ that were missed by both filters were due to the record not using the metadata employed in either filter (i.e., the search terms, subject headings, etc).

Table 5 shows whether any of the ‘includes’ missed by each filter would have been ‘Found’ or ‘Not Found’ by citation searching on Web Science or CitationChaser.

**Table 5:** Citation searching of missed ‘includes’.

| Filter: | Dataset: | Missed Record: | Web of Science |  | CitationChaser |  |
| --- | --- | --- | --- | --- | --- | --- |
|  |  |  | FCS: | BCS: | FCS: | BCS: |
| SM | 5 | **Lopez-Jaramillo et al. (1987)** | N/A | N/A | Found | Not Found |
|  | 9 | Broderick et al. (2014) | Found | Not Found | Not Found | Found |
|  | 11 | Giustino et al. (2020) | Not Found | Found | Not Found | Found |
|  | 12 | Leyburn et al. (1959) | Found | Not Found | Found | N/A |
|  | 13 | **Oertel et al. (2023)** | Found | Found | Found | Found |
|  |  | **Zinzani et al. (2016)** | Found | Not Found | Found | Not Found |
|  |  | **Rigacci et al. (2020)** | Not Found | Found | Found | Found |
| SaPM | 1 | McNamara et al. (2014) | N/A | N/A | Found | Found |
|  | 5 | **Lopez-Jaramillo et al. (1987)** | N/A | N/A | Found | Not Found |
|  |  | Raman et al. (1978) | Found | Not Found | Found | Found |
|  | 9 | Toohey et al. (2018) | Not Found | Not Found | Not Found | Not Found |
|  | 13 | **Oertel et al. (2023)** | Found | Found | Found | Found |
|  |  | **Zinzani et al. (2016)** | Found | Not Found | Found | Not Found |
|  |  | **Rigacci et al. (2020)** | Not Found | Found | Found | Found |
|  |  | Fiaccadori et al. (2023) | Not Found | Found | Not Found | Found |
|  |  | Barrington et al. (2016) | Found | Found | Found | Found |
|  |  | Eichenauer et al. (2021) | Found | Found | Found | Found |
|  |  | Kobe et al. (2018) | Found | Found | Found | Found |
|  |  | Marini et al. (2021) | Not Found | Found | Not Found | Found |
|  |  | Mettler et al. (2018) | Found | Found | Found | Found |
|  |  | Illidge et al. (2020) | Found | Found | Found | Found |
|  |  | Viviani et al. (2020) | N/A | N/A | Not Found | Found |
|  |  | Bari et al. (2020) | Found | Found | Found | Found |

**FCS** = forward citation searching; **BCS** = backward citation searching; **N/A** = not available on the citation index. **NB:** Grey cells are used to highlight records that were either not available or did not match any ‘includes’; bold text is used for records that were missed by both filters.

Using CitationChaser, all seven records missed by the SM filter and 15 of 16 records missed by SaPM filter could be retrieved if both forwards *and* backwards citation chasing was performed. For the SM filter, forwards citation searching found five of the seven missed records, with four of the seven missed records found backwards. Whereas, for the SaPM filter, forwards citation searching found 12 of 16 records and 13 of 16 records were found backwards.

Using Web of Science, six of seven records missed by the SM filter and 12 of 16 records missed by SaPM filter could be retrieved if both forwards *and* backwards citation chasing was performed. For the SM filter, forwards citation searching found four of the seven missed records, with three of the seven missed records found backwards. Whereas, for the SaPM filter, forwards citation searching found nine of 16 records and 10 of 16 records were found backwards.

Overall, Web of Science did not find any records that could not be found by searching CitationChaser (if both directions were searched), whereas CitationChaser found several records that could not be found with bidirectional citation searching on Web of Science (one for the SM filter and three for the SaPM filter).

Table 6 shows the precision of the SM and SaPM filters and number-needed-to-read for the nine datasets. The mean precision for the SM filter was 1.5% versus 4.4% by the SaPM filter, which is a difference of 2.9%. The mean NNR for the SM filter was 189 versus 68 by the SaPM filter, which is a difference of 121.

**Table 6:** Precision and Number-Needed-to-Read.

| Dataset: | Precision |  |  | NNR |  |  |
| --- | --- | --- | --- | --- | --- | --- |
|  | SM Filter | SaPM Filter | Difference in Precision: | SM Filter | SaPM Filter | Difference in NNR: |
| 1 | 2.2% | 3.3% | 1.0% | 45 | 31 | 14 |
| 2 | 1.2% | 2.1% | 0.8% | 81 | 49 | 32 |
| 4 | 0.1% | 0.4% | 0.3% | 687 | 231 | 456 |
| 8 | 0.2% | 0.6% | 0.4% | 549 | 172 | 377 |
| 9 | 1.7% | 2.4% | 0.7% | 59 | 42 | 17 |
| 10 | 2.6% | 5.4% | 2.9% | 39 | 18 | 21 |
| 12 | 1.9% | 12.1% | 10.2% | 52 | 8 | 44 |
| 13 | 2.8% | 11.2% | 8.4% | 35 | 9 | 26 |
| 14 | 0.7% | 1.8% | 1.2% | 153 | 55 | 98 |
| **Mean:** | **1.5%** | **4.4%** | **2.9%** | **189** | **68** | **121** |

**SM** = Sensitivity-Maximizing Filter; **SaPM** = Sensitivity- and Precision-Maximizing Filter.

Table 7 shows the total number of records for the reproduction search strategies with and without each filter applied, for the nine datasets. It lists the number that records were reduced by and shows this as a percentage for each filter and dataset. Differences between each filter are shown, in addition to the mean.

**Table 7:** Percentage reduction in overall number-needed-to-screen.

| Dataset: | Total Records Without Filters: | SM Filter |  | SaPM Filter |  | Difference in Reduction of Records: |
| --- | --- | --- | --- | --- | --- | --- |
|  |  | Records Reduced To (ONNS): | Records Reduced By: | Records Reduced To (ONNS): | Records Reduced By: |  |
| 1 | 1,619 | 847 | 772 (47.7%) | 551 | 1,068 (66%) | 296 (18.3%) |
| 2 | 4,506 | 2,187 | 2,319 (51.5%) | 1,317 | 3,189 (70.8%) | 870 (19.3%) |
| 4 | 31,856 | 15,807 | 16,049 (50.4%) | 5,320 | 26,536 (83.3%) | 10,487 (32.9%) |
| 8 | 17,330 | 6,583 | 10,747 (62%) | 2,058 | 15,272 (88.1%) | 4,525 (26.1%) |
| 9 | 4,212 | 1,721 | 2,491<br>(59.1%) | 1,274 | 2,938<br>(69.8%) | 447<br>(10.6%) |
| 10 | 3,757 | 1,515 | 2,242<br>(59.7%) | 716 | 3,041<br>(80.9%) | 799<br>(21.3%) |
| 12 | 1,719 | 730 | 989<br>(57.5%) | 124 | 1,595<br>(98.2%) | 606<br>(35.3%) |
| 13 | 4,813 | 1,449 | 3364<br>(69.9%) | 286 | 4,527<br>(94.1%) | 1163<br>(24.2%) |
| 14 | 2,470 | 1,833 | 637<br>(25.8%) | 664 | 1,806<br>(73.1%) | 1169<br>(47.3%) |
| **Mean:** | **8,031** | **3,630** | **4,401**<br>**(54.8%)** | **1,368** | **6,664**<br>**(83%)** | **2,262**<br>**(28.2%)** |

**SM** = Sensitivity-Maximizing Filter; **SaPM** = Sensitivity- and Precision-Maximizing Filter; **ONNS** = Overall number-needed-to-screen.

The mean ONNS for the SM filter was 3,630 with a mean record reduction of 4,401 (54.8%). The mean ONNS for the SM filter was 1,368 with a mean record reduction of 6,664 (83%). The mean difference in the reduction of records between the filters was 2,262 (28.2%).

## DISCUSSION

### Deciding which filter to use

In deciding whether to use the SaPM filter over the SM filter, we might consider: is there a potential risk of missing eligible records; is it necessary to reduce the screening burden, and, if so, by how much; and can the potential risk of missing eligible records be mitigated in a way that reduces the screening burden? This study has found that there would generally have been low risk of missing ‘included’ records if the SaPM filter had been chosen over the SM filter for the datasets tested. Overall, the SM filter retrieved 387 of 394 (98.2%) MEDLINE ‘includes’ versus 378 of 394 (95.9%) by the SaPM filter, which is a difference of 2.3% or nine records.

In terms of recall, the SaPM filter’s performance was equal to or better than the SM filter for 11 of the 14 datasets. Dataset 13 had the highest number of ‘includes’ missed for a single dataset: 12 by the SaPM filter, versus three by the SM filter. This dataset is an outlier in the results shown in Table 3, without which the filters appear similar for recall and unique yield. This may have been caused by multiple factors. Firstly, the use of Cooper’s phase III extension filter in the search strategies for the original Cochrane review could have caused ‘includes’ to be missed by both filters. Secondly, the topic of the review (drugs) is also notable, since the SaPM filter does not search for drug therapy as a floating subheading and this metadata was in several (n=5) of its missed records.

Despite its reduced sensitivity, the SaPM filter can have advantages over the SM filter. Out of a total of seven records missed by the SM filter, three of these (42.9%) were retrieved with the SaPM filter. The three records missed by the SM filter and not the SaPM filter were due to ‘trial’ in the title field (n=3) and ‘Clinical Trials as Topic’ as a subject heading (n=1). Using the term ‘trial’ in the title of the study is recommended in the Consolidated Standards of Reporting Trials (CONSORT) guidance [27], highlighting that adaptations to the SM filter could enhance sensitivity. Characteristics of the three records uniquely found by the SaPM filter are noted in Table 4. Notably, two out of three (66.7%) had no abstract. As the SaPM filter is unique in searching the title field, it is less reliant on a record having an abstract. Records less likely to have an abstract on MEDLINE could include publication types such as errata, retractions, preprints [28] and older records.

Differences in design between the filters means their performance in comparison to each other may vary by review topic. For instance, as the SM filter has a line for drug therapy as a floating subheading, this filter could be more valuable for reviews looking to retrieve records on drugs, which could be explored in a further study. Out of a total of 16 records missed by the SaPM filter, 12 of these were retrieved with the SM filter. These 12 records missed by the SaPM filter and not the SM filter were due to the inclusion of: ‘trial’ in the abstract field (n=8); ‘drug therapy’ as a floating subheading (n=5); and ‘groups’ in the abstract field (n=2). All five records that used ‘drug therapy’ as a floating subheading came from dataset 13, which highlights the risk of using the SaPM filter in the context of this specific review. Information specialists looking to reduce the screening burden on reviews focussed on drugs as a topic or intervention could consider adapting the SaPM filter to include the floating subheading for drug therapy to enhance sensitivity. This also suggests the value of making study filter choices (or adaptations) based on the specific focus of the review.

The SaPM filter’s risk of missing eligible records could be mitigated with bidirectional citation searching. Although it can be beneficial to use multiple tools during the citation searching process, CitationChaser was the best tool to use for citation searching using the data from this study. Overall, all seven records missed by the SM filter and 15 of 16 records missed by SaPM filter were retrieved using CitationChaser. This means there was no advantage to using both Web of Science and CitationChaser, to retrieve the records missed by either filter. Table 5 shows the importance of citation searching both forwards and backwards, as many records could be found in one direction only. This data should be understood in the context of the methodology, which was to use the missed records to perform citation searching, rather than the final ‘includes’ of a review. Therefore, anything found backwards with the missed ‘includes’ would have been found forwards with the final ‘includes’ (and vice versa).

The SaPM filter had a mean precision of 4.4% versus 1.5% by the SM filter, which is a difference of 2.9%. The higher precision of the SaPM filter led to its mean NNR of 68, versus 189 by the SM filter. This figure could be used to estimate potential time and cost savings of using the SaPM filter over the SM filter, where use of the SaPM filter is considered appropriate.

On average there was an additional reduction of 2,262 records (28.2%) in ONNS when using the SaPM filter over the SM filter. However, any records retrieved by the SM filter and not by the SaPM filter might also be retrieved by database searches of other sources, which could negate the reduction in the screening burden. Therefore, our exploration of the reduced screening burden is limited as only one database was looked at in isolation.

Although both filters are widely used in search strategies for evidence syntheses, many information specialists employ additional changes or extension filters to enhance sensitivity. Notably, Cooper et al’s 2019 study found both the SM and SaPM filters might miss studies that refer to the trial phase and not the study design or randomization method [29]. Cooper et al’s phase III (P3) filter can therefore be used as an extension to either filter [29]. As shown in Table 2, other changes to the SM filter made by information specialists include adapting randomized to randomi?ed, using the wildcard ‘?’ to retrieve spelling variants; changing the field codes searched; and including additional lines for RCTs. Information specialists looking to increase sensitivity could also adapt the SM filter to include the unique lines of the SaPM filter as represented in Table 1 or examine the reasons records were missed by the SM filter (see Table 4) and consider including some of these terms.

Validated RCT filters designed for other databases are also recommended in the *Cochrane Handbook*, including filters for Ovid Embase and Embase.com [30]. However, there is not currently a filter designed to maximize both sensitivity and precision for Embase, which could be addressed in future research.

### Implications of machine learning on limiting records to RCTs

It is worth considering whether the introduction of machine-indexed subject headings and publication types on MEDLINE from April 2022 [31] could affect the ongoing performance of the Cochrane RCT filters. Notably, Askin et al reported that machine indexing of human studies was nearly twice as likely to be incorrect versus manual indexing [32], which has implications for the use of the MEDLINE limit to remove animal studies within both the SM and SaPM filters (though this was not included in our comparison). As various other studies by Guo et al [33], Smalheiser et al [34], Chen et al [35], and Amar-Zifkin et al [36] have also questioned the accuracy of machine-indexed subject headings on MEDLINE, this is an area which warrants further investigation – both in relation to search filters and the RCT filters specifically.

As an alternative to the use of RCT study filters, machine learning classifiers can also limit records to RCTs, and can be employed on systematic review software such as EPPI-Reviewer [37]. On EPPI-Reviewer, this categorises records as ‘possibly an RCT’ or ‘probably not an RCT’ and allocates a percentage range indicating how accurate this classification is likely to be [37]. Cochrane advise that RCT classifiers are best used with search strategies that do not already limit records by this study type [3]. Therefore, as an RCT classifier can be applied to records across all databases and sources, this circumvents the issue of selecting a filter for each database.

Although the design of study classifiers may be less transparent in comparison with search filters, a 2021 study by Thomas et al on EPPI-Reviewer’s RCT classifier reported 0.99 for recall and 0.08 for precision [38]; this is higher sensitivity but lower precision than the 2008 version of the SM and SaPM filters achieved in Glanville et al’s 2020 study. The higher sensitivity of the RCT classifier may suggest a larger overall number of records to screen. However, the categorisation of records in terms of how likely they are to be RCTs allows review teams to identify genuine RCTs sooner. The option of RCT classifiers, especially in combination with the use of machine-learning to prioritise screening (which re-orders the screening list by relevance based on screening decisions) [37], could therefore be a valuable option for review teams with an unmanageable number of hits or with limited resources.

## LIMITATIONS

This is an exploratory study in which only 394 MEDLINE records taken from healthcare literature searches were tested. While this focus on only 14 Cochrane reviews may not be sufficiently broad to indicate general trends, it provides insight into choosing between Cochrane’s updated RCT filters for Ovid MEDLINE and explores whether any risks of missing eligible records could be mitigated with citation searching.

The study’s methodology, which used the final ‘includes’ of completed Cochrane reviews that had used the SM filter, does not provide a true comparison between the filters. However, it is valuable to understand the SaPM filter’s performance in the context of the SM filter, which tends to be used as standard. Additionally, as all 14 Cochrane reviews used the 2008 versions of the filters, the ‘includes’ found may not be representative of the records that could have been found had the 2023 revision been used. However, this does not affect the comparison of the 2023 updated filters in relation to each other, as they were both updated in the same way. This data also allows us to understand differences between the updated filters in terms of their precision, NNR and ONNS for the selected Cochrane reviews.

We did not include the lines to remove animal studies within each filter, as information specialists may not always employ this limit. Use of the limit would therefore have generated different results for the precision, NNR, on ONNS reduction for each filter.

Searching for studies using all or part of the title of the record could have introduced minor errors. However, all records were double-checked.

Citation searching was only conducted using two programs. Moreover, as it was performed in 2025, the results may not reflect those available to review teams at the time. A further limitation is that the percentage reduction in ONNS examines MEDLINE in isolation, so any reduction in records might be negated by other database searches returning some of these ‘reduced’ records.

## CONCLUSION

Although information specialists might expect that the SaPM filter cannot retrieve any unique records versus the SM filter, this is incorrect. Of the 394 records tested in our study, the SM filter had a unique yield of 12, versus three unique records found by the SaPM filter. This is important data to consider in deciding between the filters, which could necessitate minor adaptations to certain lines in the filter of choice, depending on the needs and topic of the review.

This study found that using the SaPM filter for Ovid MEDLINE could reduce title and abstract screening by an additional 28.2% on average over the SM filter, whilst retrieving 95.9% of ‘includes’ compared with 98.2% with the SM filter. The SaPM filter offered an 83% reduction in ONNS compared with 54.8% by the SM filter. Moreover, the SaPM’s average NNR was 68, versus 189 for the SM filter. Use of the SaPM filter could save time for larger review topics or projects with shorter deadlines. Moreover, the potential risk of missing eligible records could be mitigated by citation searching. Further research could test how the filters compare in retrieval of drug studies, since the SaPM filter missed several drug studies that were retrieved by the SM filter. It would also be valuable to consider how the filters compare to the RCT study classifier on EPPI-Reviewer and to establish the accuracy of records tagged with the subject headings used in both filters since the introduction of automated indexing in April 2022 [31].

Overall, our findings are informative to help information specialists understand the performance of the SaPM filter in the context of the performance of the SM filter on Ovid MEDLINE. In deciding whether it is appropriate to use the SaPM filter over the SM filter, review teams could consider the trade-off between potentially missing a small number of eligible records and the efficiency savings made, which might enhance other aspects of the review. The filters should be carefully compared on a review-by-review basis and consider citation searching to mitigate the risk of missing eligible records.

## ACKNOWLEDGEMENTS

We would like to acknowledge NLM Support for their time and help answering our enquiries. We are also grateful to all the peer reviewers for their diligence and helpful feedback.

## CONFLICT OF INTEREST STATEMENT

The authors have no competing interests to report.

## SOURCES OF FUNDING STATEMENT

The authors have no sources of funding to report.

## ETHICS STATEMENT

Ethical approval was not necessary for this study.

## PEER REVIEW STATEMENT

This study was submitted to a journal in July 2025 and has undergone a single round of peer reviews.

## DATA AVAILABILITY STATEMENT

Data associated with this article are available in the appendix and can be obtained by contacting the first author.

## Notes

### Competing Interest Statement

The authors have declared no competing interest.

## REFERENCES

[1] Salvador-Oliván JA, Marco-Cuenca G, Arquero-Avilés A. Development of an efficient search filter to retrieve systematic reviews from PubMed. Journal of the Medical Library Association. 2021;109(4):561–574. 10.5195/jmla.2021.1223

[2] Glanville JM, Lefebvre C, Miles J N, & Camosso-Stefinovic J. How to identify randomized controlled trials in MEDLINE: ten years on. Journal of the Medical Library Association. 2006;94(2):130–136.

[3] Lefebvre C, Glanville J, Briscoe S, Featherstone R, Littlewood A, Metzendorf M-I, Noel-Storr A, Paynter R, Rader T, Thomas J, Wieland LS. Chapter 4: Searching for and selecting studies [last updated March 2025]. In: Higgins JPT, Thomas J, Chandler J, Cumpston M, Li T, Page MJ, Welch VA, editors. 2025. Cochrane Handbook for Systematic Reviews of Interventions, version 6.5.1 https://training.cochrane.org/handbook/current/chapter-04

[4] Lefebvre C, Glanville J, Briscoe S, Featherstone R, Littlewood A, Metzendorf M-I, Noel-Storr A, Paynter R, Rader T, Thomas J, Wieland LS. Technical Supplement to Chapter 4: Searching for and selecting studies [last updated September 2024]. In: Higgins JPT, Thomas J, Chandler J, Cumpston M, Li T, Page MJ, Welch VA, editors. 2024. Cochrane Handbook for Systematic Reviews of Interventions, version 6.5. cochrane.org/handbook

[5] Glanville J, Kotas E, Featherstone R, & Dooley G. Which are the most sensitive search filters to identify randomized controlled trials in MEDLINE? Journal of the Medical Library Association. 2020;108(4):556–563. 10.5195/jmla.2020.912

[6] Personal communication [Internet]. E-mail from NLM Support. 9 May 2025.

[7] Sampson M, Tetzlaff J, & Urquhart C. Precision of healthcare systematic review searches in a cross-sectional sample. Research Synthesis Methods. 2011;2:119–125. 10.1002/jrsm.42

[8] Bethel AC, Rogers M, & Abbott R. Use of a search summary table to improve systematic review search methods, results, and efficiency. Journal of the Medical Library Association. 2021;109(1):97–106. 10.5195/jmla.2021.809

[9] Cooper C, Garside R, Varley-Campbell J, Talens-Bou J, Booth A, Britten N. “It has no meaning to me.” How do researchers understand the effectiveness of literature searches? A qualitative analysis and preliminary typology of understandings. Research Synthesis Methods. 2020;11:627–640. 10.1002/jrsm.1426

[10] Frequently Asked Questions about Indexing for MEDLINE [Internet]. National Library of Medicine; n.d. [cited 25 April 2025]. https://www.nlm.nih.gov/bsd/indexfaq.html

[11] Sampson M, Zhang L, & Morrison A, et al. An alternative to the hand searching gold standard: validating methodological search filters using relative recall. BMC Medical Research Methodology. 2006;6:33. 10.1186/1471-2288-6-33

[12] Campisi SC, Zasowski C, Bradley-Ridout G, Schumacher A, Szatmari P, & Korczak D. Omega-3 fatty acid supplementation for depression in children and adolescents. Cochrane Database of Systematic Reviews. 2024. 10.1002/14651858.CD014803.pub2

[13] Ernst M, Wagner C, Oeser A, Messer S, Wender A, Cryns N, et al. Resistance training for fatigue in people with cancer. Cochrane Database of Systematic Reviews. 2024. 10.1002/14651858.CD015518

[14] Esezobor CI, Bhatt GC, Effa EE, Hodson EM. Fenoldopam for preventing and treating acute kidney injury. Cochrane Database of Systematic Reviews. 2024. 10.1002/14651858.CD012905.pub2

[15] Kolkailah AA, Abdelghaffar B, Elshafeey F, Magdy R, Kamel M, Abuelnaga Y, et al. Standard-versus extended-duration anticoagulation for primary venous thromboembolism prophylaxis in acutely ill medical patients. Cochrane Database of Systematic Reviews. 2024. 10.1002/14651858.CD014541.pub2

[16] Kongwattanakul K, Duangkum C, Ngamjarus C, Lumbiganon P, Cuthbert A, Weeks J, Sothornwit J. Calcium supplementation (other than for preventing or treating hypertension) for improving pregnancy and infant outcomes. Cochrane Database of Systematic Reviews. 2024. 10.1002/14651858.CD007079.pub4

[17] Sachdeva A, Rai BP, Veeratterapillay R, Harding C, & Nambiar A. Non-steroidal anti-inflammatory drugs for treating symptomatic uncomplicated urinary tract infections in non-pregnant adult women. Cochrane Database of Systematic Reviews. 2024. 10.1002/14651858.CD014762.pub2

[18] Ferraro MC, Urquhart DM, Ferreira GE, Wewege MA, Abdel Shaheed C, Traeger AC, et al. Antidepressants for low back pain and spine-related leg pain. Cochrane Database of Systematic Reviews. 2025. 10.1002/14651858.CD001703.pub4

[19] Reid J, Blair C, Dempster M, McKeaveney C, Slee A, Fitzsimons D. Multimodal interventions for cachexia management. Cochrane Database of Systematic Reviews. 2025. 10.1002/14651858.CD015749.pub2

[20] Wagner C, Ernst M, Cryns N, Oeser A, Messer S, Wender A, et al. Cardiovascular training for fatigue in people with cancer. Cochrane Database of Systematic Reviews. 2025. 10.1002/14651858.CD015517

[21] Chan M, Chan JJ, Wright JM. Effect of amphetamines on blood pressure. Cochrane Database of Systematic Reviews. 2025. 10.1002/14651858.CD007896.pub4

[22] Al Said S, Kaier K, Nury E, Alsaid D, Gibson CM, Bax J, et al. Non-vitamin K antagonist oral anticoagulants (NOACs) after transcatheter aortic valve replacement (TAVR): a network meta-analysis. Cochrane Database of Systematic Reviews. 2025. 10.1002/14651858.CD013745.pub2

[23] Spillane J, Trip J, Drost G, Faber CG, Hanna MG, Nevitt SJ, Vivekanandam V. Drug treatment for myotonia. Cochrane Database of Systematic Reviews. 2025. 10.1002/14651858.CD004762.pub3

[24] Kreuzberger N, Goldkuhle M, von Tresckow B, Kobe C, Sickinger MT, Monsef I, Skoetz N. Positron emission tomography-adapted therapy for first-line treatment in adults with Hodgkin lymphoma. Cochrane Database of Systematic Reviews. 2025. 10.1002/14651858.CD010533.pub3

[25] Wang GM, Li LJ, Fan L, Xu M, Tang WL, Wright JM. Renin inhibitors versus angiotensin receptor blockers for primary hypertension. Cochrane Database of Systematic Reviews. 2025. 10.1002/14651858.CD012570.pub2

[26] Briscoe S, Bethel A, & Rogers M. Conduct and reporting of citation searching in Cochrane systematic reviews: A cross-sectional study. Research Synthesis Methods. 2020;11(2):169–180. 10.1002/jrsm.1355

[27] Schulz KF, Altman DG, Moher D, & CONSORT Group. CONSORT 2010 Statement: updated guidelines for reporting parallel group randomised trials. BMC Medicine. 2010;8(18):1–9. 10.1186/1741-7015-8-18

[28] PubMed User Guide [Internet]. National Library of Medicine; 2025. [cited 23 May 2025]. https://pubmed.ncbi.nlm.nih.gov/help/

[29] Cooper C, Varley-Campbell J, & Carter P. Established search filters may miss studies when identifying randomized controlled trials. Journal of Clinical Epidemiology. 2019;112:12–19. 10.1016/j.jclinepi.2019.04.002

[30] Glanville J, Foxlee R, Wisniewski S, Noel-Storr A, Edwards M, & Dooley G. Translating the Cochrane EMBASE RCT filter from the Ovid interface to Embase.com: a case study. Health Information and Libraries Journal. 2019;36(3), 264–277. 10.1111/hir.12269

[31] Use of MeSH in Indexing [Internet]. National Library of Medicine; n.d. [cited 25 April 2025]. https://www.nlm.nih.gov/mesh/intro_indexing.html

[32] Askin N, Ostapyk T, Epp C. Filtering failure: the impact of automated indexing in Medline on retrieval of human studies for knowledge synthesis. Journal of the Medical Library Association. 2025;113:58–64. 10.5195/jmla.2025.1972

[33] Guo M, Gotz D, Wang Y. How does imperfect automatic indexing affect semantic search performance? [Internet]. arXiv; 2023 [cited 2023 Apr 15]. http://arxiv.org/abs/2304.04057

[34] Smalheiser NR, Shahidehpour A, Troy AM. Issues regarding the indexing of adaptive clinical trial articles. medRxiv: the preprint server for health sciences. 2025. 10.1101/2025.03.10.25323694

[35] Chen E, Bullard J, Giustini D. Automated indexing using NLM’s Medical Text Indexer (MTI) compared to human indexing in Medline: a pilot study. Journal of the Medical Library Association. 2023;111(3):684–94. 10.5195/jmla.2023.1588

[36] Amar-Zifkin A, Ekmekjian T, Paquet V, & Landry T. Algorithmic indexing in MEDLINE frequently overlooks important concepts and may compromise literature search results. Journal of the Medical Library Association. 2025;113(1):39–48. 10.5195/jmla.2025.1936

[37] Machine learning functionality in EPPI-Reviewer [Internet]. EPPI Centre, Social Science Research Unit, Institute of Education, University of London; n.d. [cited 25 April 2025]. https://eppi.ioe.ac.uk/CMS/Portals/35/Machine%20learning%20in%20EPPI-Reviewer%20v_10%20web%20version.pdf

[38] Thomas J, McDonald S, Noel-Storr A, et al. Machine learning reduced workload with minimal risk of missing studies: development and evaluation of a randomized controlled trial classifier for Cochrane Reviews. Journal of Clinical Epidemiology. 2021;133:140–151. 10.1016/j.jclinepi.2020.11.003

